# Positive rates predict death rates of Covid-19 locally and worldwide 13 days ahead

**DOI:** 10.1101/2020.11.24.20237842

**Authors:** Jürgen Mimkes, Rainer Janssen

## Abstract

In the Covid-19-pandemic, the numbers of deceased do not consistently follow the number of new infections. The CFR mortality has declined in Germany from 5 % to 0.4 %. However, if we interpret the portion of positive tests as a positive rate, we find the positive rate and the numbers of deceased to run parallel with an offset of about 13 days. This has been observed worldwide in ten other countries and locally in Germany and North Rhine-Westphalia. In Germany the IFR mortality is about 29 per one million inhabitants, in the USA about 42, in Israel about 17, in the Netherlands 23, in Austria 27, in France 33, in Spain 36, in the UK 47 and Italy about 56 per million inhabitants. In Japan and South-Korea the mortality rate is only about 3 per million inhabitants, with an offset of about 25 days.

The daily positive ratio, which is reported by state health authorities, allows to estimate the number of deaths (and seriously ill people) about 13 days ahead. This gives local hospitals more time for detailed planning. The daily positive rate may be interpreted as a “thermometer” of the respective country. The positive rate gives a much better picture of the state of the pandemic and should be reported by the media in addition to infection numbers. In official guidelines a 7-day-positive-rate is a much better guideline than the 7-day-incidence.

## Introduction

Mortality is the main indicator of the fatality of the COVID-19 pandemic [1-3]. According to the World Health Organization (WHO), there are two ways to determine mortality:

### *IFR Mortality* (infection fatality ratio)

The numbers of deceased follow the number of all newly infected by a fraction IFR and a time lap L,

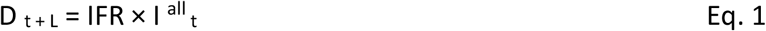

D _t + L_ represent the numbers of deceased on day (t + L), and I ^all^ _t_ is the number of all people infected on day (t). In the COVID-19-pandemic, however, many infected people show few or no symptoms and are not recorded. This makes IFR mortality difficult to apply.

### *CFR Mortality* (case fatality ratio)

In practice mortality is calculated from the number of confirmed cases and the fatality ratio CFR,

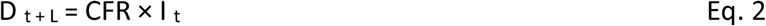

D _t + L_ represent the numbers of deceased on day (t + L), I _t_ is the number of cases confirmed on day (t).

## Data

For scientific analysis of Covid-19, mortality data sets for many countries are available, containing the numbers of daily deaths, daily new infections and daily test volumes [4 - 7]. The following two figures show Covid-19 pandemic data for Germany.

Fig. 1 shows the daily number of confirmed new infections and fig. 2 the daily number of deaths in Germany. The data sets for newly infected and for deceased are shifted against each other by about 13 days, a small fraction of the infected died on average after about L = 13 days.

**Fig. 1.**
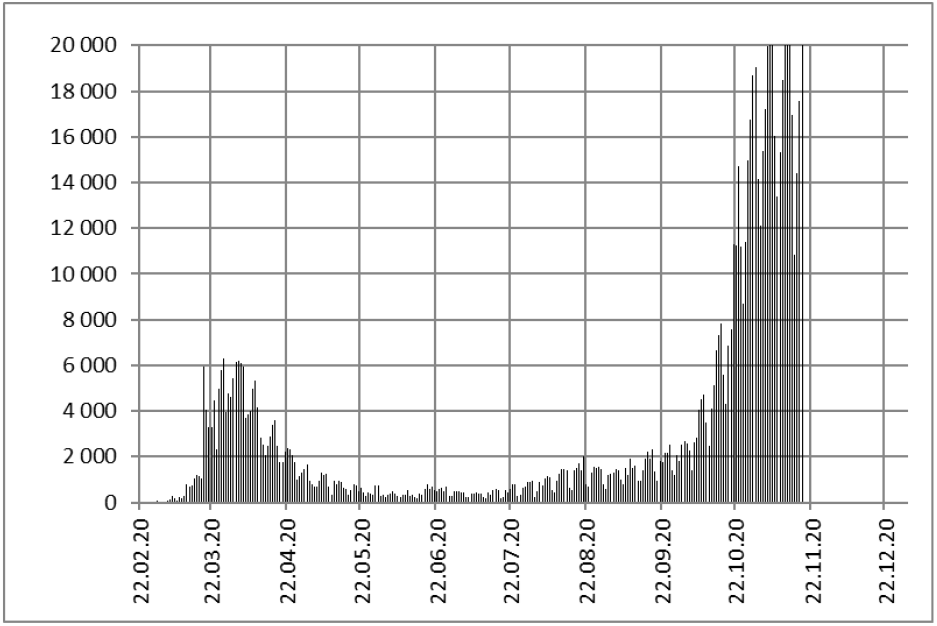
Confirmed daily infections in Germany [5]

**Figure. 2.**
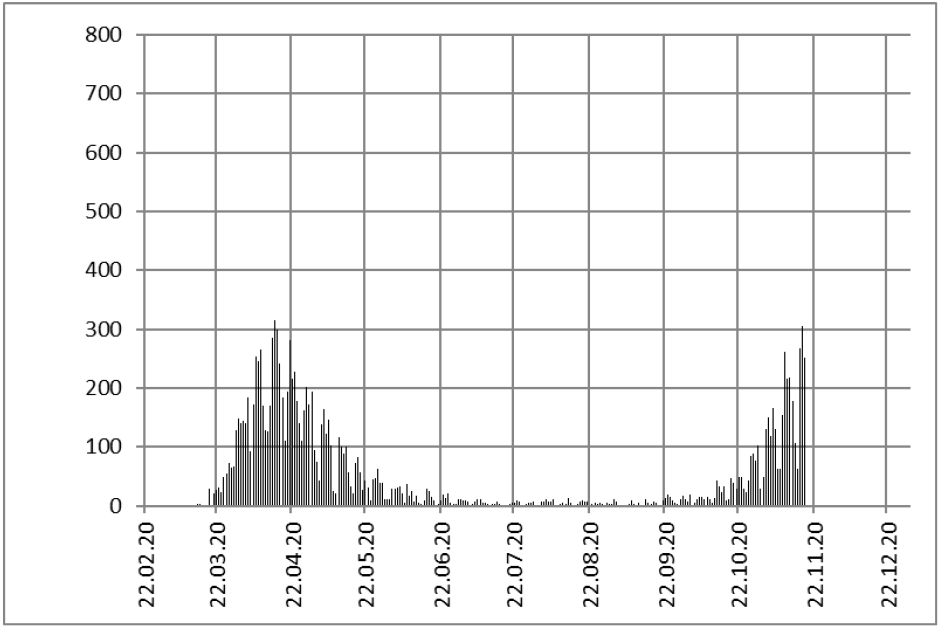
Daily number of deaths in Germany [5]

However, the two series in fig. 1 and 2 are not proportional, as would be expected according to eq. 2. CFR mortality is not constant in Germany, but has decreased during the pandemic from CFR = 5 % to 0,4 % and has increased again, later on [3]. The decline of mortality has been explained by the Robert Koch Institute (RKI), providing evidence for a time-depending age shift of patients suffering from Covid-19. While in the first wave mainly elderly people were tested and fell ill, the median age of the sick fell significantly in July.

Nevertheless, a time variation in CFR mortality may also indicate a fundamental difficulty in obtaining mortality.

### Positive rate of tests

As many infected patients of Covid-19 do not show symptoms, many infections do not show up as cases. However, if we interpret daily infections I _t_ as positive tests of a test volume T _t_, we can define the daily positive rate P _t_ of tests by

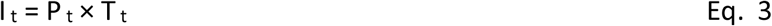

The positive rate P _t_ of tests may be regarded as a concentration of infections in a population.

We may compare this with the concentration of salt in seawater. In order to determine the salt concentration, it is not necessary to measure the whole sea, it is sufficient to take samples. By evaporating 1 liter of seawater, one will find 33 g of salt, evaporating 2 liters will give 66 g of salt. But 66 g salt does not make sense without the test volume, and 20 000 new infections become only meaningful by specifying the test volume.

### IFR Mortality of Covid-19

We may now find the law of IFR mortality by expanding the tests to the entire population, T _t_ = N. Now we obtain from eq. 3 the total number of infections: I ^all^ _t_ = N × P _t_, and we may calculate the IFR mortality according to eq. 1,

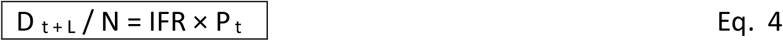

In eq. 4 the calculated relative number of daily deceased D _t + L_ / N is proportional to positive rate P _t_. The proportional factor is the IFR mortality, N is the size of the population.

One can observe the similarity between the number of deaths in fig. 2 and the positive rate P _t_ in fig. 3. We have confirmed the similarity of figures 2 and 3 by calculating the numbers of deceased according to eq. 4 and plotting the result in fig. 4. The numbers of daily deaths (vertical bars) and calculations according to the IFR mortality in eq. 4 (solid line) agree well, in contrast to the CFR mortality calculations of eq. 2 given as dotted line.

**Fig. 3.**
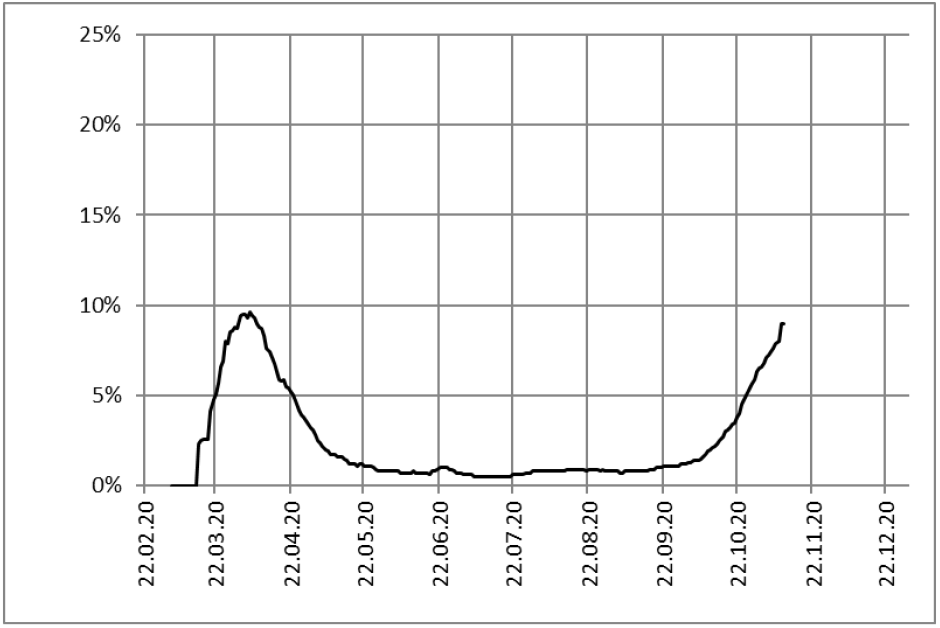
Positive rate [6]

**Fig. 4.**
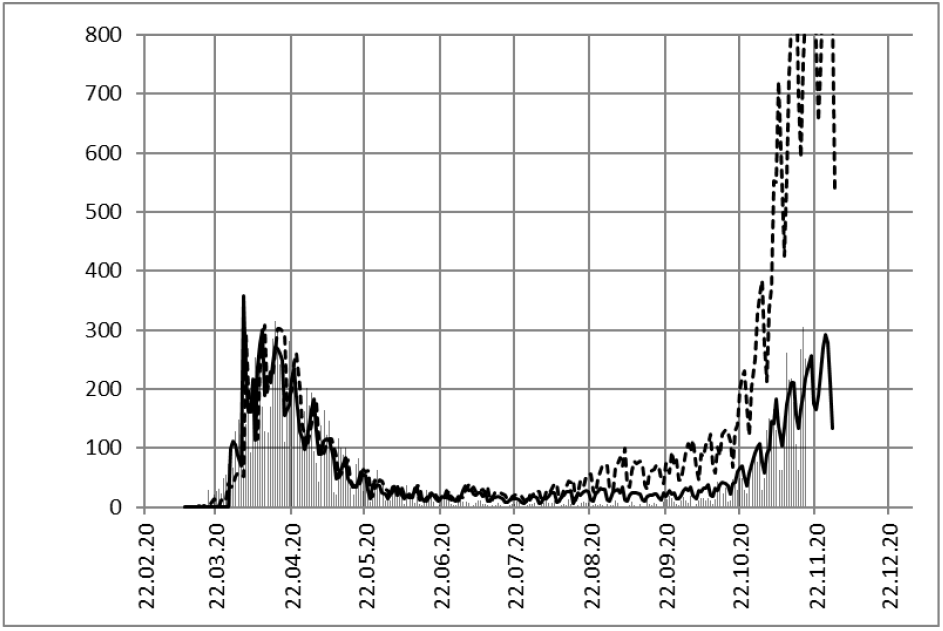
Actual daily deaths (vertical bars), mortality calculations according to Eq. 4 (line) and Eq. 2 (dashed)

The factor of mortality in Germany is determined by scaling the calculations to the data [8; 9],

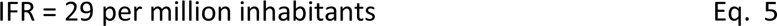

For example, in the week from 12.10.20, the positive rate averaged 2.6 %. According to eq. 4, the number of deaths per week from 25.10.20 (i.e. 13 days later) is:

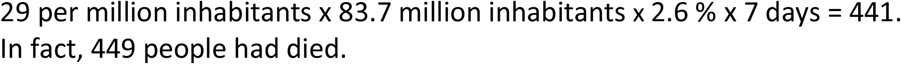

The estimation of deaths during summer shows a deviation which may be investigated later.

### Estimating the total number of deaths

Eq. 4 also refers to the total number of deaths if all daily deaths are summed up. This summation requires daily positive rates of the state or county. The state of North Rhine-Westphalia (NRW) in Germany shows about the same positive rate as Germany in total [10]. For the district of Paderborn, Table 1 assumes the same positive rate as for NRW, as positive rates were not available for the county. In table 1 the number of deaths has been calculated according to eq. 4 and compared to the actual data.

**Table 1.** Number of all deceased persons calculated for 21/11/20

| | Inhabitants/Mill. | Sum $P_t$ | deaths calculated | deaths registered |
| --- | --- | --- | --- | --- |
| **Germany** | 84 | 5.86 | **14 224** | 13 884 |
| **NRW** | 18 | 5.86 | **3 058** | 2 943 |
| **Paderborn** | 0.3 | 5.86 | **51** | 56 |

The agreement between Germany, NRW and the district of Paderborn shows that the IFR mortality is about the same all over the country. This estimate can be made by any other state or county, but the daily positive rates have to be available.

### Estimating the death numbers in Germany 13 days ahead

The number of deaths follows the number of new infected persons after about 13 days. Accordingly, we may calculate the number of deaths 13 days ahead. Fig. 5 shows the calculations according to eq. 4.

**Fig. 5.**
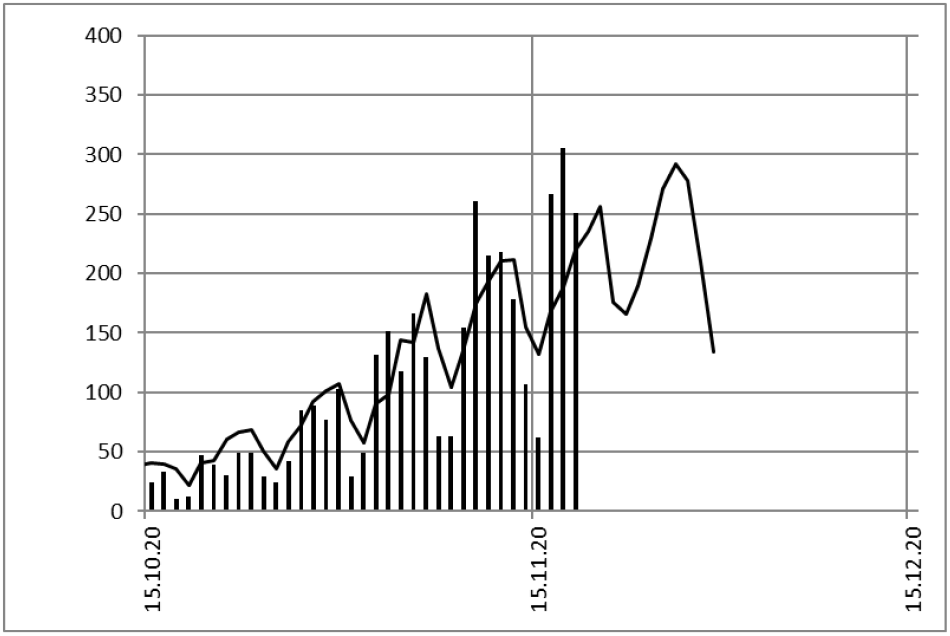
Deceased (vertical bars) and estimation of deaths by Eq. 4 (Line) in Germany 13 days ahead

In fig. 5 the estimate is based on the IFR of eq. 5. The higher death numbers at the end may be due to fluctuations, but could also indicate a rise of the IFR in Germany in the second wave. This will be clarified later.

The estimate of death numbers 13 days in advance can also be carried out for states, federal states or districts, if the daily positive rates are known. Unfortunately, there are often no positive rates for counties. But this would be very important, as not the state but the hospitals of the county have to be prepared for the pandemic.

### The “thermometer” of the Covid-19-pandemic

The positive rate in eq. 4 may be interpreted as the “thermometer” of the Covid-19 pandemic. Until now, the authorities and media have always used the numbers of new infections as an indicator of the pandemic. Johns Hopkins University ranks countries by the total number of infections. The 7-day incidence also refers to the number of new infections to justify official measures such as closing schools or restaurants or a lockdown to bring down the number of patients in the hospitals.

A better indicator of the epidemic would actually be the numbers of deceased, which demonstrates the danger and does not depend on test numbers. The large number of deaths in Italy at the beginning of the pandemic has convinced many Germans of the danger of the new pandemic. But according to eq. 4, this “thermometer” is 13 days late. It shows the situation two weeks ago and is less suitable as an instruction to act.

The real “thermometer” of the COVID-19 pandemic is the positive rate in eq. 4. It is proportional to the numbers of deceased, but indicates the danger immediately and not 13 days later. With this positive rate it is possible to estimate the daily number of deaths and the number of patients who have to go to a hospital.

Modelling the development of the pandemic should also refer not to confirmed infections, but to the positive rate in order to make the model independent of the test volume.

### Positive rate and IFR mortality in different countries

Figures 6 and 7 show positive rates and deaths in the US and the calculation by IFR and CFR mortality. Again, we observe the consistency between the positive rate and the actual number of deaths.

**Figure 6.**
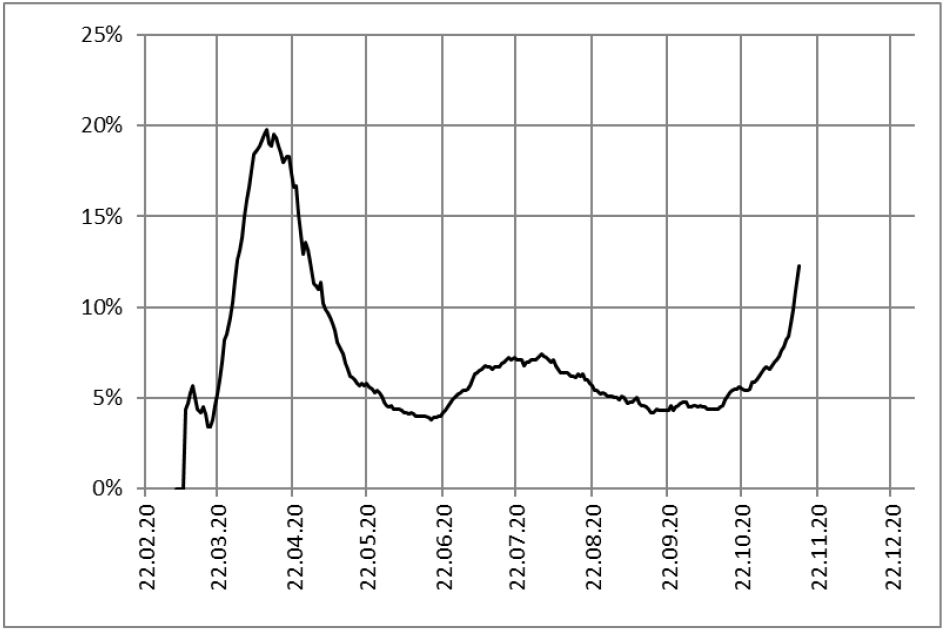
Positive rate [6] in USA

**Fig. 7.**
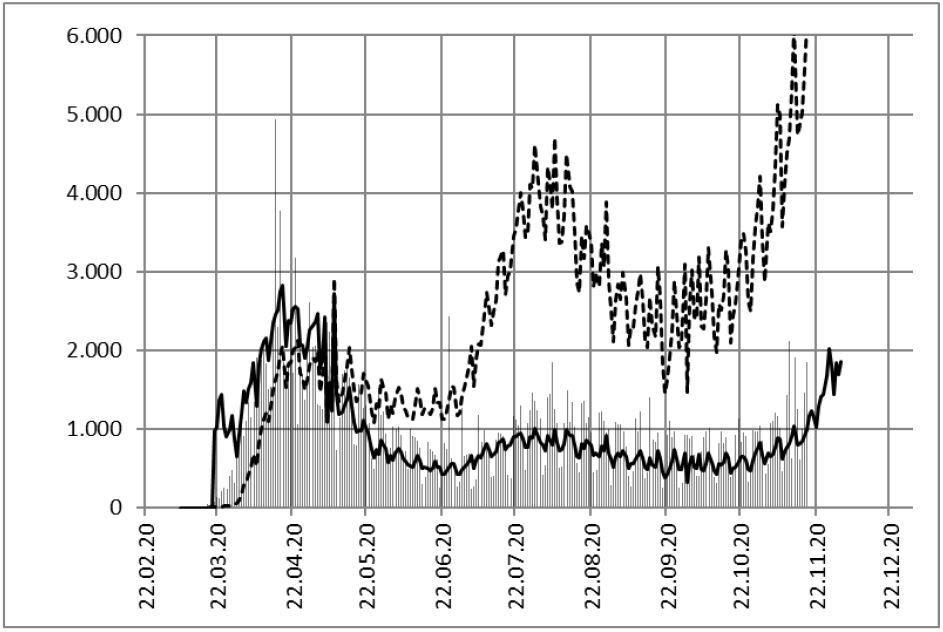
Actual daily deaths (vertical bars), deaths by IFR (line) and by mortality CFR (dotted) mortality in USA

The IFR mortality rate in the USA is about 42 per million inhabitants.

Figures 8 to 16 show calculations of the number of deaths according to eq. 4 compared to the data [6] for each country. The resulting IFR factors are in UK 47, in France 33, in BRA 29, in Israel 17, in Italy 56. In the Netherlands we obtained 23, in Austria 27 and in Spain 36 per million inhabitants, however, for these countries the data set for the positive ratio was not complete. In Japan and South Korea, on the other hand, the mortality rate is only about 3 per million inhabitants, with a latency period of about 25 days.

**Fig. 8.**
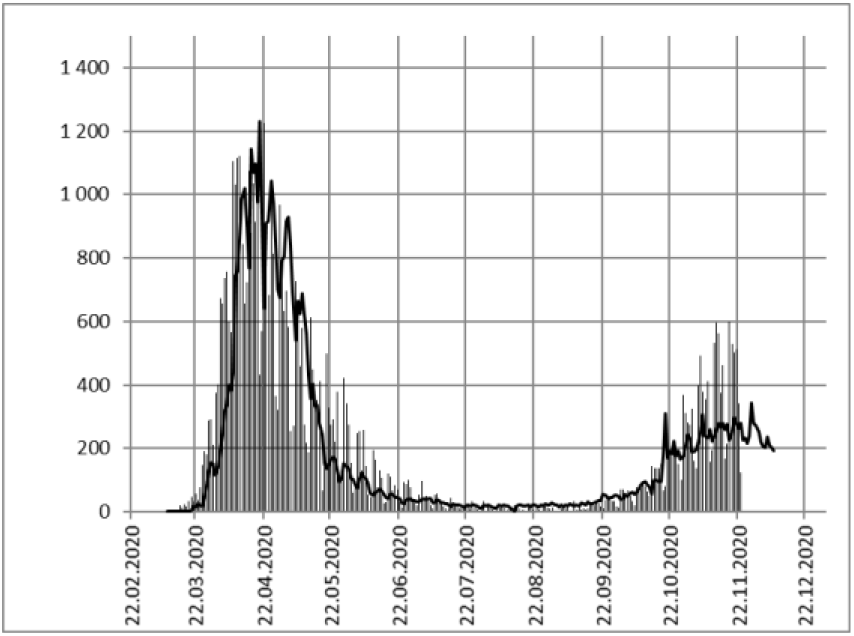
**UK:** IFR = 47 per million inhabitants

**Fig. 9.**
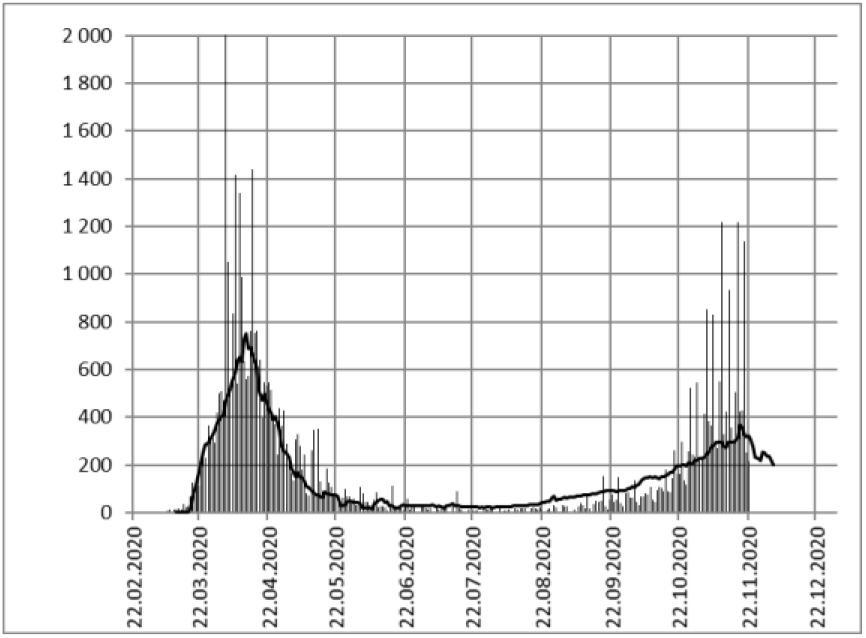
**FRA:** IFR = 33 per million inhabitants

**Fig. 10.**
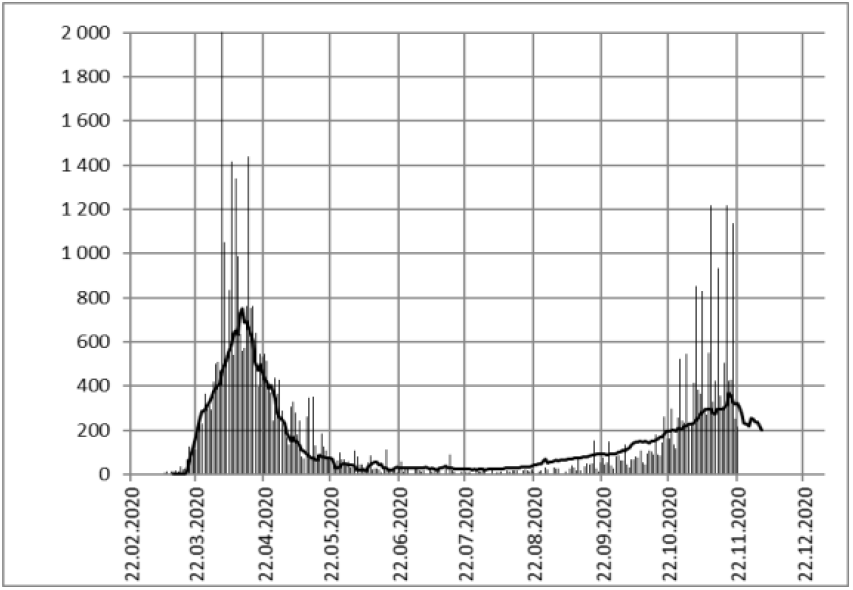
**BRA:** IFR = 29 per million inhabitants

**Fig. 11.**
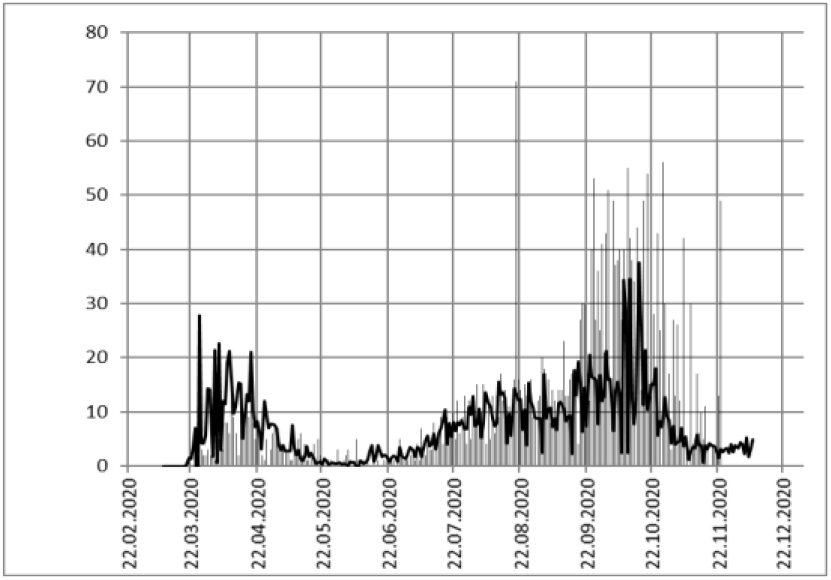
**ISR:** IFR = 17 per million inhabitants

**Fig. 12.**
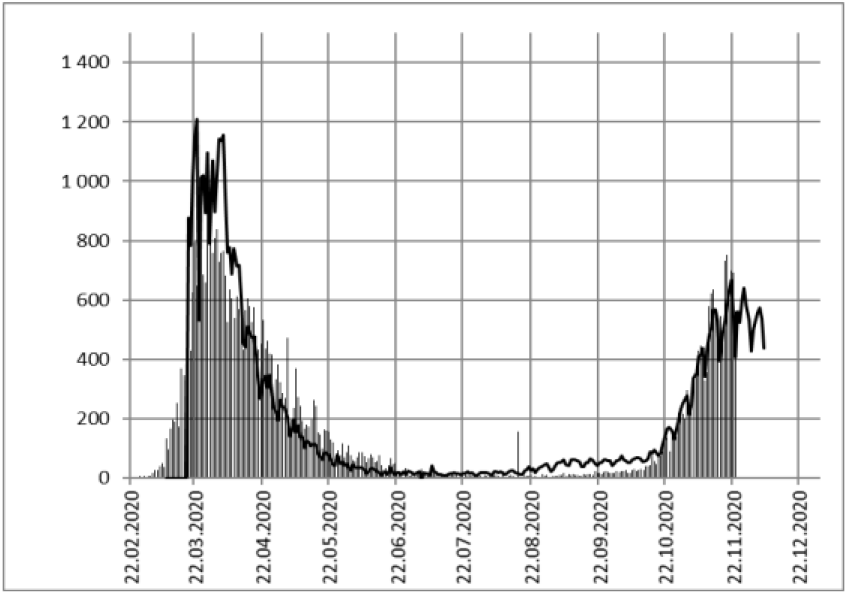
**ITA:** IFR = 56 per million inhabitants

**Fig. 13.**
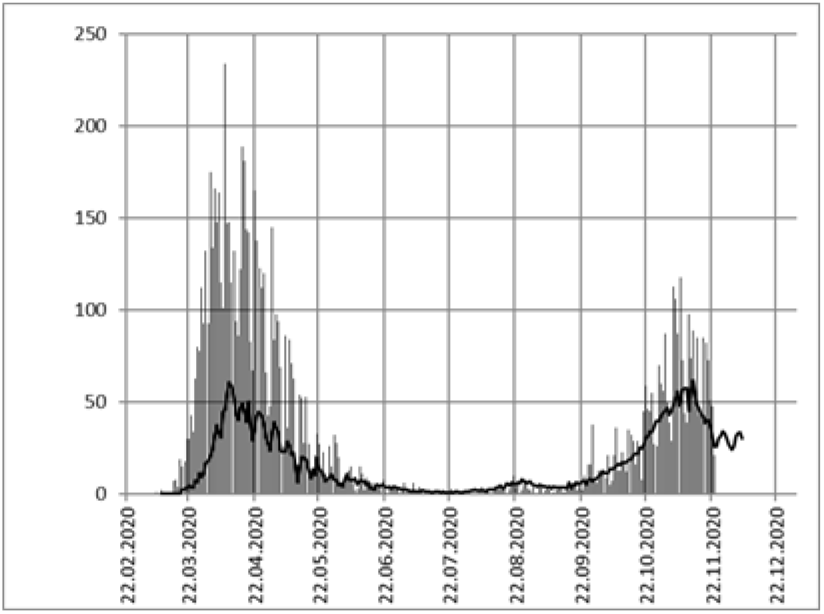
**NLD:** IFR = 23 per million inhabitants

**Fig. 15.**
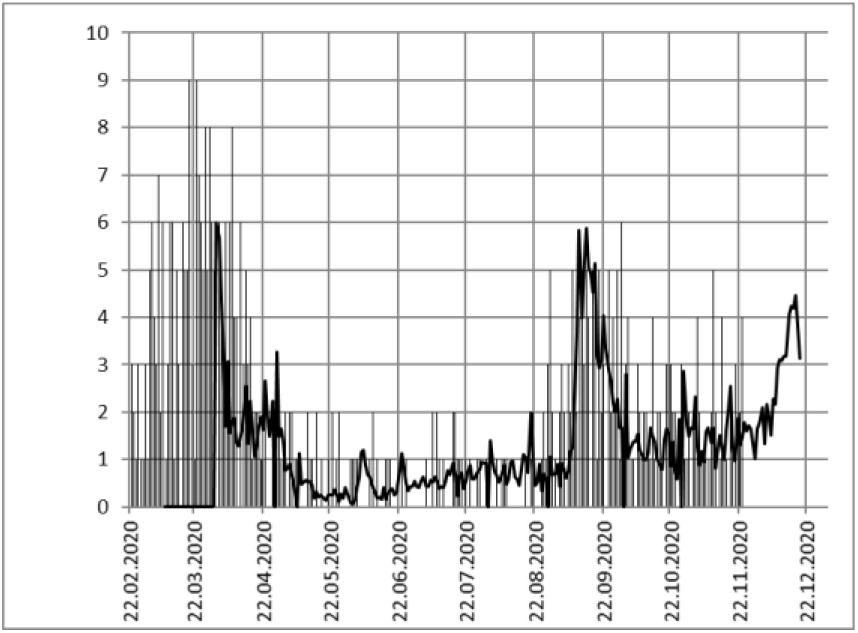
**KOR:** IFR = 3 per million inhabitants

**Fig. 16.**
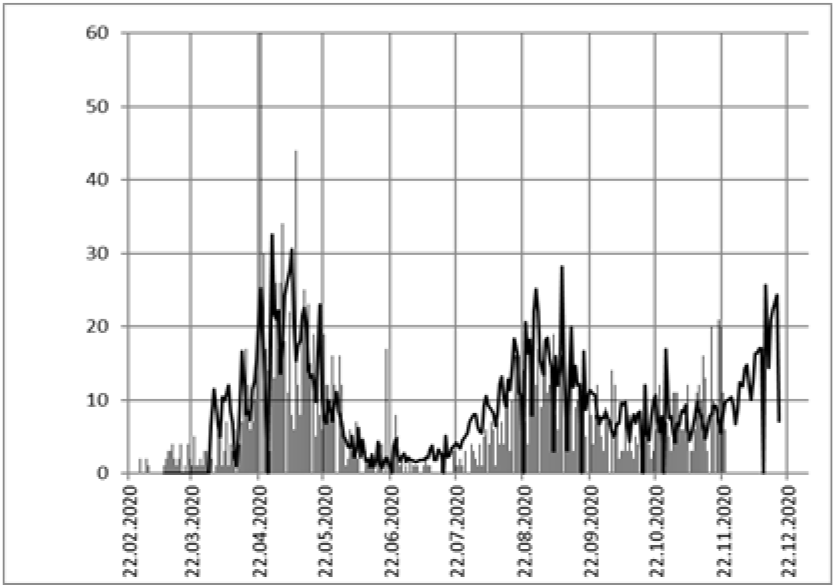
**JPN**: IFR = 3 per million inhabitants

## Conclusions

The numbers of new infections have dominated the headlines since the beginning of the Covid-19 pandemic. But the positive rate should be supplemented as soon as possible by media and officials. Only the positive rate can be used to compare the risk level of the pandemic in different states or countries. Medical institutions can estimate the number of deaths and seriously ill patients two weeks ahead.

**Number of daily deaths and calculation of IFR mortality in eight additional countries**

## Data Availability

All data are available on servers open to public

https://physik.uni-paderborn.de/alumni/mimkes

